# Global Burden of Covid-19 Restrictions: National, Regional and Global Estimates

**DOI:** 10.1101/2021.09.21.21263825

**Authors:** Günther Fink, Fabrizio Tediosi, Stefan Felder

**Affiliations:** Swiss Tropical and Public Health Institute; University of Basel; Faculty of Business and Economics, University of Basel

**Author notes:** Corresponding author Günther Fink.

## Abstract

A large literature has documented the high global mortality and mental health burden associated with the current Covid-19 pandemic. In this paper, we combine newly collected data on subjective reductions in the quality of life with the latest data on Covid-19 restrictions to quantify the total number of quality-adjusted life years (QALYs) lost due to government imposed restrictions globally. Our estimates suggest a total loss of 2980 (95% 2764, 3198) million QALYs as of September 6^th^ 2021, with the highest burden absolute burden in lower and upper middle income countries. QALY losses appear to be particularly large for closures of schools and daycares as well as restaurants and bars, and seem relatively small for wearing masks in public and closure of fitness facilities.

## Main

The COVID-19 pandemic has resulted in unprecedented social, economic and health systems disruptions globally. According to the latest estimates, 20.5 million years of life have been lost to COVID-19 to date,^1^ and millions of new cases continue to be recorded each week despite the rollout of vaccines in many countries and the continued use of masks as well as other preventive measures in most settings.^2^

Measures to reduce the spread of Covid-19 have been of paramount importance to avoid major health system breakdowns and to limit excess mortality during peak infection periods as those seen in Northern Italy in April of 2020^3^ or in India approximately one year later.^4^ While these measures are widely considered a success from an epidemiological and public health perspective,^5,6^ they also have come at a substantial cost for governments. The direct economic cost of Covid-19 measures have been estimated at USD 7.7 trillions for the US alone^7^ and have resulted in unprecedented increases in government debt in many countries.^8,9^

A large number of studies has attempted to assess the cost-effectiveness of measures imposed to restrict Covid-19 transmission.^10^ Most of the existing cost-effectiveness assessments either compare estimated life years gained to the financial cost of measures faced by governments,^11,12^ or estimate the relative cost per life year saved for different containment strategies.^13^ Both approaches essentially abstract from the loss in the quality of life experienced by individuals and families because of these restrictions, including lost early life learning opportunities^14^, limited access to schooling, loss of employment and, in some cases, complete social isolation. The impact of these restrictions are partially visible in the increased incidence of loneliness,^15^ increased prevalence of mental health problems both among adolescents^16^ and adults^17^ as well as a general deterioration in living conditions, particularly in low income settings.^18^ However, the reductions in general well-being go well beyond these specific dimensions of well-being. Life under Covid-19 restrictions entails not only lack of personal and physical contact and frequent social isolation, but also having to combine home office work with child care duties, being deprived of access to sports and entertainment facilities, and frequently also not getting access to specialized medical services.

For decision making in health, the overall well-being of individuals or patients with specific health conditions or restrictions is generally established through standardized surveys that quantify the subjective valuation of specific states relative to a (healthy) life without these conditions. In standardized quality of life surveys, states are defined over a specific health condition such as blindness or paraplegia, and survey respondents asked to indicate how much they value life with this condition relative to a fully healthy life through a series of time tradeoff questions (TTOs).^19^ These responses and relative valuations can then be used to quantify the quality-adjusted life years (QALYs) lost due to a specific condition or the QALY gains of treatment.

### Quality of life under Covid-19 restrictions

To assess the relative well-being of the population under Covid-19 restrictions, we asked survey respondents in France, India, Italy, the UK and the US to complete a series of time tradeoff question related to living with light restrictions (masks, restricted access to restaurants, restricted international travel) and severe restrictions (light restrictions plus home office, home schooling and restrictions to private gatherings). We also asked respondents to specify the relative utility of life with paraplegia as a commonly used reference point. A total of 952 persons completed the survey across the five countries. The average estimated utility was 0.71 (95% CIs 0.69-0.74) for light restrictions, 0.65 (0.63-0.68) for severe restrictions, and 0.49 (0.47-0.51) for paraplegia. As shown in **Figure 1**, QALY utility weights were relatively similar across countries (Figure 1). Lowest average utility weights were found in France, and highest weights on average in India. Highest disutility was found for the 40-49 age group and lowest disutilities for individuals 70+. For gender, no differences were found overall, but patterns varied quite substantially across countries (Figure 2b). Supplemental Materials Figure 2 shows the full empirical distribution of (individual-level) relative utilities by country.

**Figure 1:**
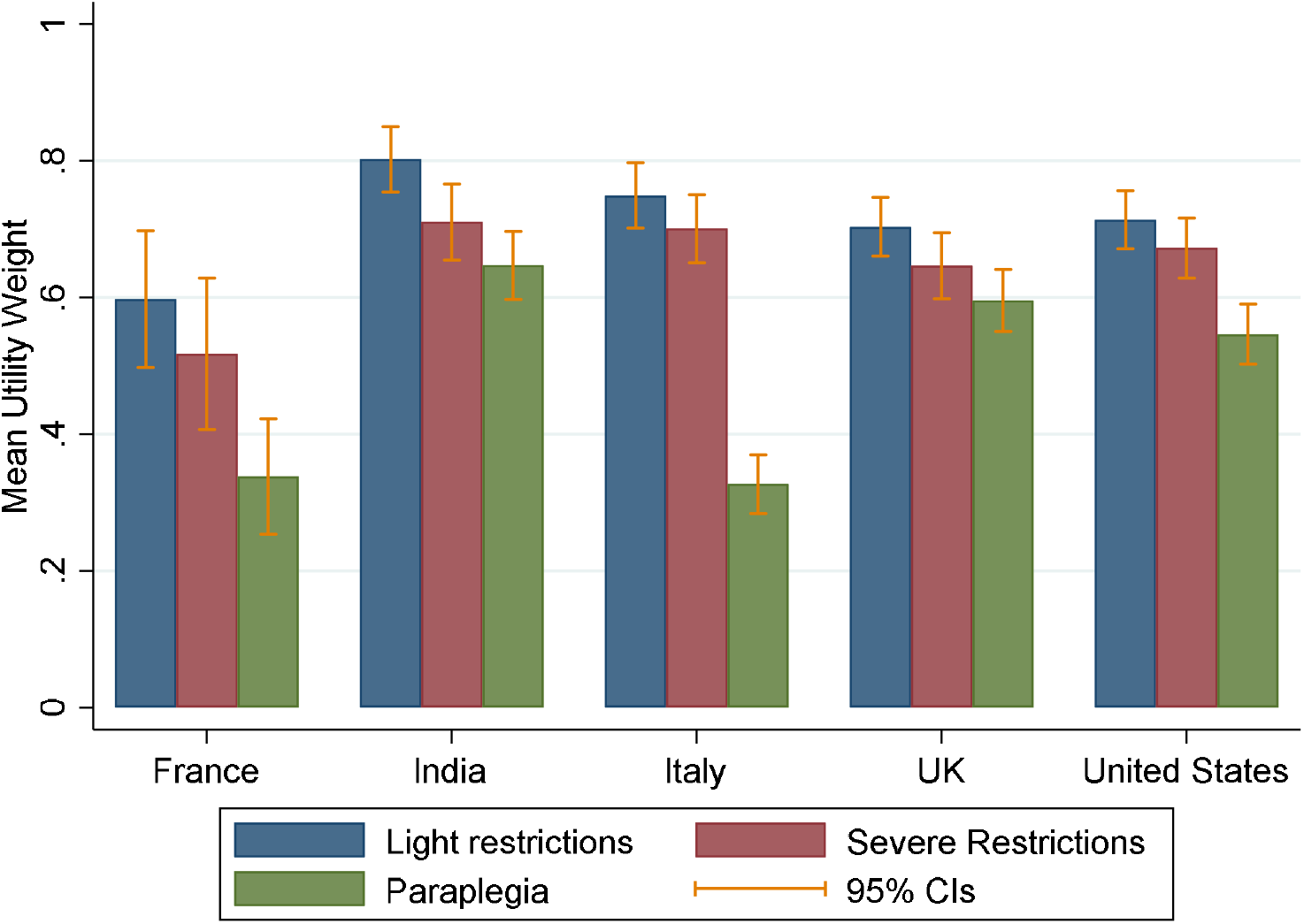
Mean utility weights for light restrictions, severe restrictions and paraplegia. *Notes:* Light restrictions: wearing masks in public spaces, restricted access to bars and restaurants, limited international travels. Severe restrictions: wearing masks in public spaces, restricted access to bars and restaurants, limited international travels. Mandatory home office, remote schooling and the inability to hold private meetings.

**Figure 2:**
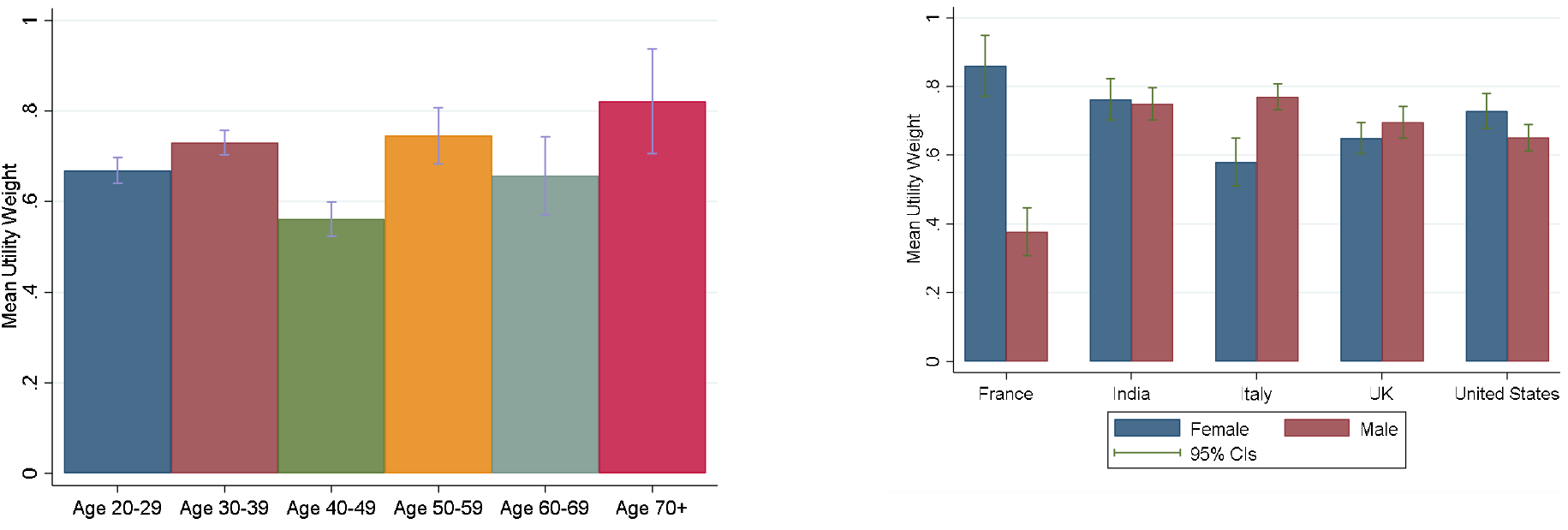
Stratified Utility Weights. Notes: Figure 2 shows estimated average utility weights by age and gender. Age estimates are based on the weighted pooled sample and include both light and severe restrictions. Gender estimates were computed separately for each country.

### Total QALYs lost due to Covid-19 restrictions

Figure 3 summarizes the extent of Covid-19 related restrictions up to 6^th^ September 2021 as compiled by the Oxford Covid-19 Governmental Response Tracker.^2^ On average, countries experienced 4.6 months of light (Stringency Index between 20-60) and 11.1 month of severe restrictions (Index over 60) between January 1, 2020 and September 6, 2021. The two countries with fewest restrictions to date were Burundi and Nicaragua; the countries with the longest severe restrictions were Azerbaijan and Canada. Supplemental Materials Figures S3 and S4 provide separate country-level maps for light and severe restrictions.

**Figure 3:**
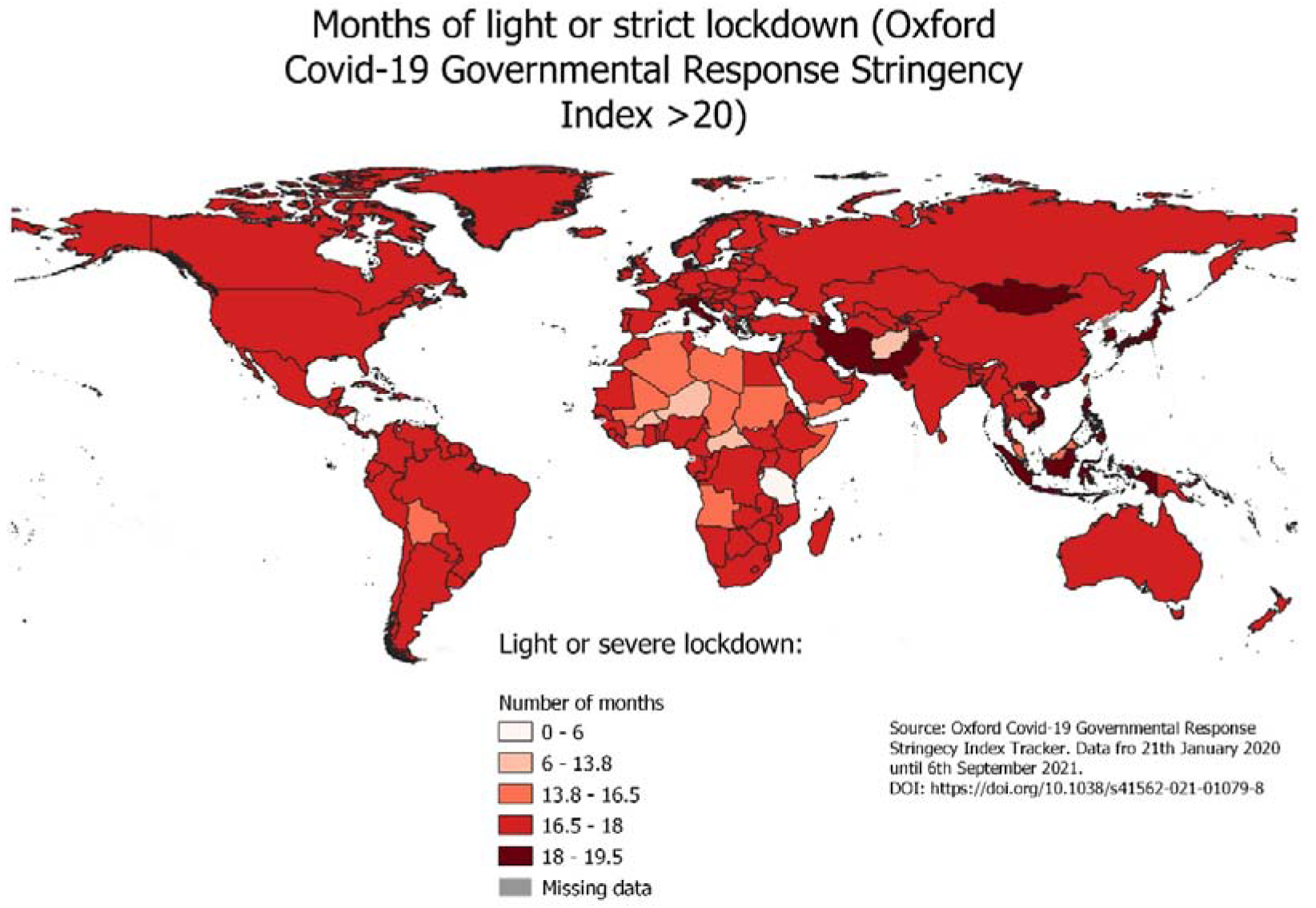
Months of light or severe restrictions by country between Jan 21, 2020 and Sept 6, 2021.

Globally, an estimated total of 2980 million QALYs (95% CIs 2764, 3198) have been lost to date due to light or severe restrictions **(Table 1)**; the majority of this burden is concentrated in upper and lower middle income countries due to their large populations as well as long average duration of restrictions.

**Table 1:**
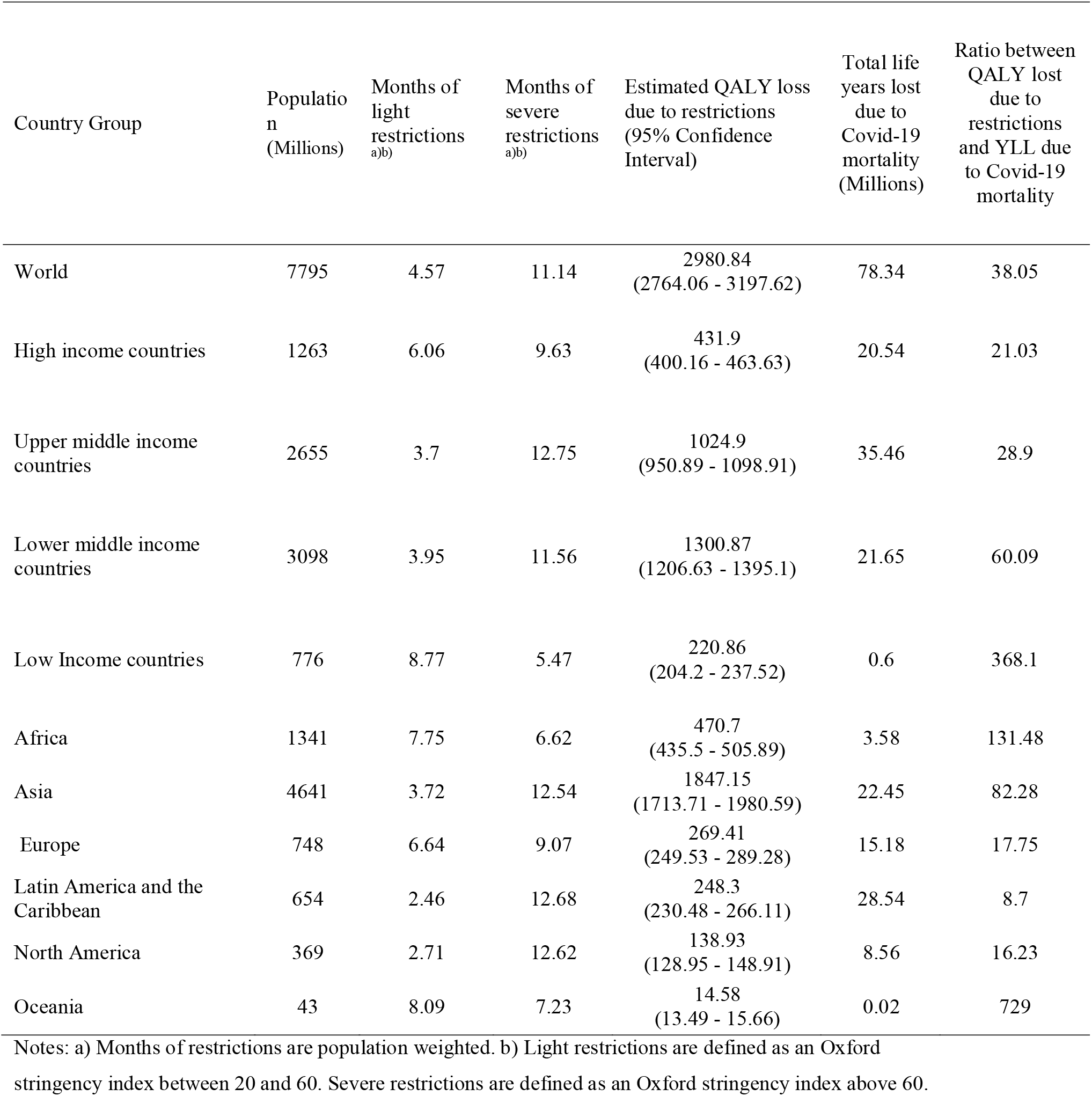
Total QALYs lost due to restrictions as well as YLLs due to Covid-19 mortality.

| Country Group | Population<br>(Millions) | Months of<br>light<br>restrictions<br>a)b) | Months of<br>severe<br>restrictions<br>a)b) | Estimated QALY loss<br>due to restrictions<br>(95% Confidence<br>Interval) | Total life<br>years lost<br>due to<br>Covid-19<br>mortality<br>(Millions) | Ratio between<br>QALY lost<br>due to<br>restrictions<br>and YLL due<br>to Covid-19<br>mortality |
| --- | --- | --- | --- | --- | --- | --- |
| World | 7795 | 4.57 | 11.14 | 2980.84<br>(2764.06 - 3197.62) | 78.34 | 38.05 |
| High income countries | 1263 | 6.06 | 9.63 | 431.9<br>(400.16 - 463.63) | 20.54 | 21.03 |
| Upper middle income<br>countries | 2655 | 3.7 | 12.75 | 1024.9<br>(950.89 - 1098.91) | 35.46 | 28.9 |
| Lower middle income<br>countries | 3098 | 3.95 | 11.56 | 1300.87<br>(1206.63 - 1395.1) | 21.65 | 60.09 |
| Low Income countries | 776 | 8.77 | 5.47 | 220.86<br>(204.2 - 237.52) | 0.6 | 368.1 |
| Africa | 1341 | 7.75 | 6.62 | 470.7<br>(435.5 - 505.89) | 3.58 | 131.48 |
| Asia | 4641 | 3.72 | 12.54 | 1847.15<br>(1713.71 - 1980.59) | 22.45 | 82.28 |
| Europe | 748 | 6.64 | 9.07 | 269.41<br>(249.53 - 289.28) | 15.18 | 17.75 |
| Latin America and the<br>Caribbean | 654 | 2.46 | 12.68 | 248.3<br>(230.48 - 266.11) | 28.54 | 8.7 |
| North America | 369 | 2.71 | 12.62 | 138.93<br>(128.95 - 148.91) | 8.56 | 16.23 |
| Oceania | 43 | 8.09 | 7.23 | 14.58<br>(13.49 - 15.66) | 0.02 | 729 |

Notes: a) Months of restrictions are population weighted. b) Light restrictions are defined as an Oxford stringency index between 20 and 60. Severe restrictions are defined as an Oxford stringency index above 60.

Figure 4 illustrates the ratio of QALYs lost due to restrictions to life years lost due to Covid-19 mortality. On average, the ratio between restriction-driven QALY losses and YLLs due to Covid-19 was 38:1, with particularly high ratios in Asia and Africa.

**Figure 4:**
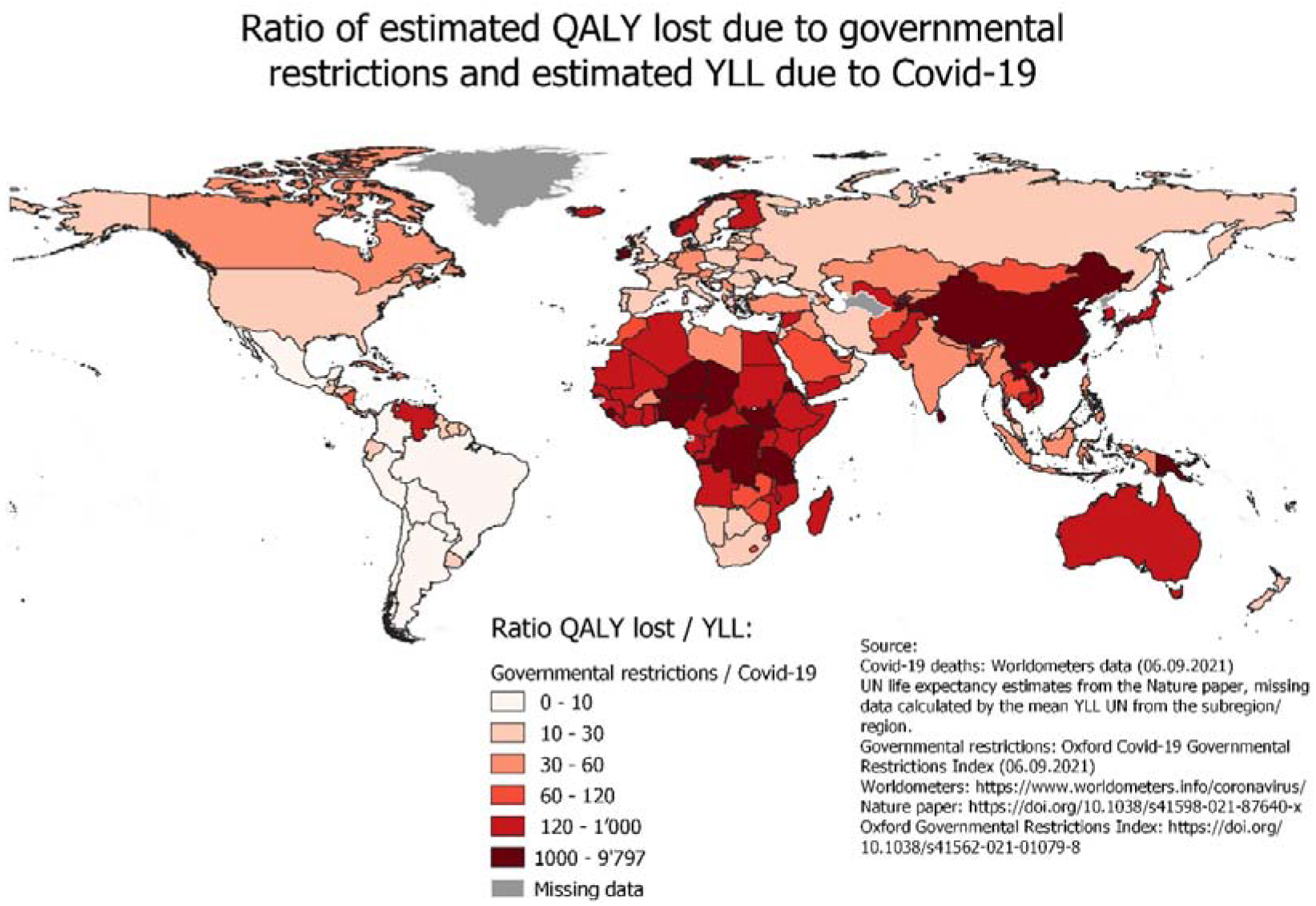
Ratio of QALYs lost due to Covid-19 Restrictions to YLL due to Covid-19.

### Willingness to pay (WTP) to avoid restrictions

In order to quantify the respondents’ WTP to avoid specific restrictions, we invited all study participants to also participate in a discrete choice experiment (DCE). As part of this DCE, we asked subjects to choose between bundles of living conditions involving restrictions on everyday life as well as pre-specified incomes. We then used random utility models to estimate (implicit) valuations of each restriction. **Figure 5** shows that across all countries, subjects were willing to give up 22% (95% CI 0.18-0.27) of their annual salary to avoid school closures and willing to give up 21.8% (95% CI 0.17-0.26) to avoid closures of restaurants, bars and clubs. Lowest WTP was observed for removing travel restrictions (7%, 95% CI 0.02-0.12) and wearing masks in public (2% (95% CI -0.02, 0.07). Full details on the specific questions asked as well as the choice sets given to study participants are provided in the Supplemental Materials below.

**Figure 5:**
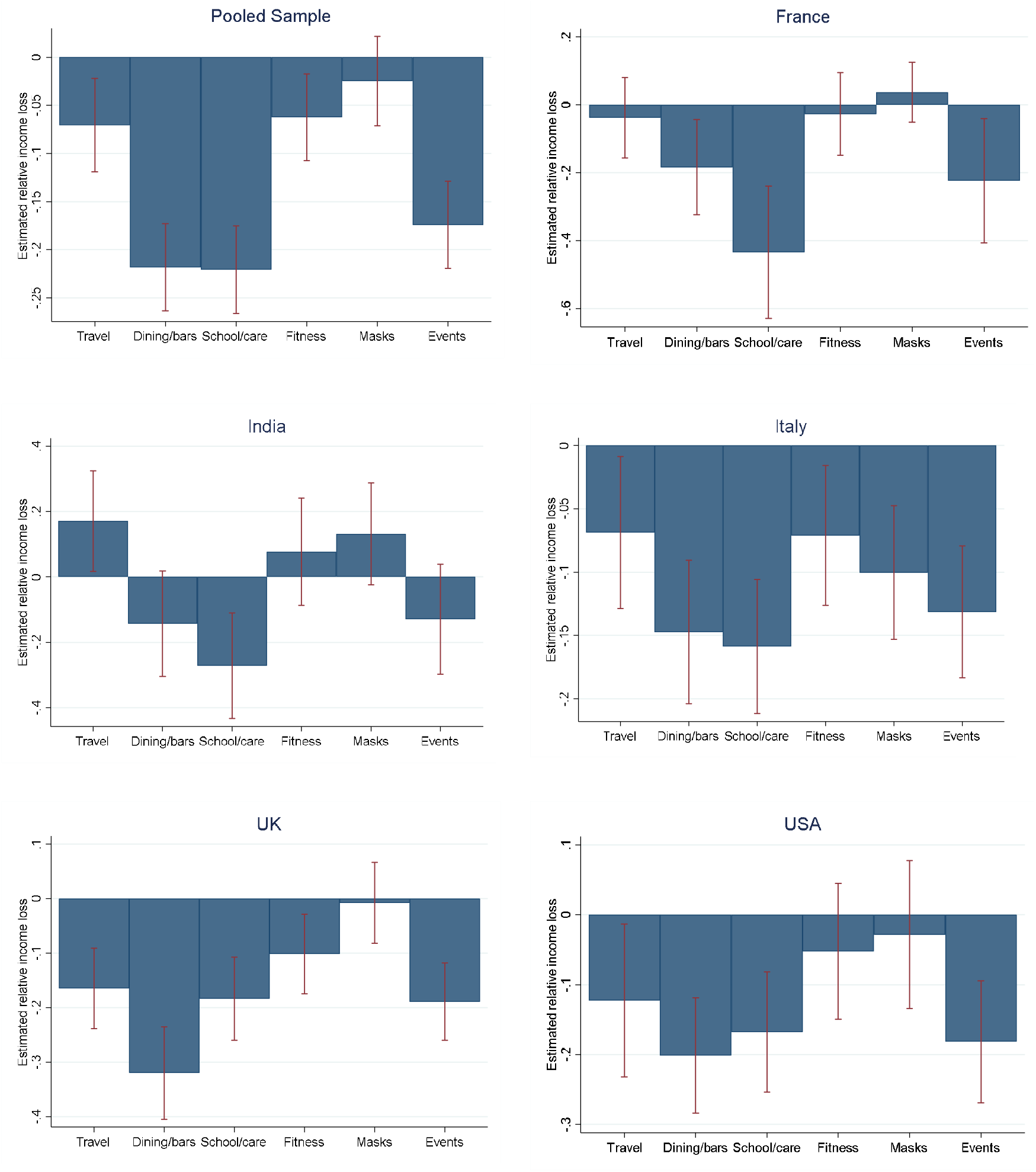
Estimated WTP per year (% of income) for avoiding specific restrictions. *Notes*: Figures show estimated WTP for avoiding each restriction as a proportion of incomes. A relative income loss of -0.1 implies that on average respondents are willing to give up 10% of their incomes to avoid the specific measure. Estimates are based on random utility logistic regression. For France and Italy, the median monthly salary used was Euro 2000. For India, the UK and the US, median annual salaries used as reference point were RP 260,000, UKP 30,000 and USD 50,000, respectively.

## Discussion

Despite the impressive progress made with respect to Covid-19 vaccinations in many high income countries, non-pharmaceutical interventions remain a key, and in many low income countries the main strategy to control new outbreaks of Covid-19. In this paper, we show that the societal burden of these measures amounts to almost three billion QALYs as of September 6, 2021, which corresponds to 38 times the estimated number of life years lost due to the epidemic so far. Even though the ratio of QALYs to mortality life years lost is rather striking, it does not imply that measures taken so far were excessive or inappropriate: without any doubt, mortality would have been much higher in many countries without the measures taken, and health systems would be in much worse conditions today. However, our results do strongly suggest that the societal costs of any restrictive measures taken by governments may be larger than what is commonly acknowledged, and that most citizens would likely be willing to give up a substantial fraction of their incomes to avoid several of these measures in the future. These large costs need to be carefully considered when developing new strategies to contain viral spread over the coming months and years. While some measures like wearing masks in public spaces or restrictions on international travels are perceived to be only a minor burden by most study participants and can still be quite effective in reducing disease transmission (Abaluck et al, 2021), the individual and social losses due to other measures such as closures of schools and the closure of bars and restaurants are substantial. As data on the relative effectiveness of specific measures becomes increasingly available based on the global experience with the first three waves, effectiveness estimates should be carefully weighed against the financial and population-level impacts of each restriction in the next phase of the epidemic.

Even though this study is to our knowledge the first attempt to quantify the societal impact of Covid-19 restrictions at both the national and global level, several limitations are worth highlighting. First, we were only able to collect survey data in five countries. Even though we found only relatively small differences in the stated utility weights across these somewhat diverse countries, it is possible that larger differences in the subjective valuation of measures would be found in a larger or more diverse sets of countries. Second, while we used census-based sampling weights to create nationally representative samples, it is also possible that respondents may not be fully representative of their respective age, gender, educational attainment stratum. Empirically, the differences across age, gender and educational attainment groups seem relatively small on average, which suggests that minor changes in sample composition will likely only have very small effects on the overall QALY losses estimated. The third limitation of the study is that there are currently no internationally validated questionnaires to estimate QALY utilities for states that are only indirectly health related such as Covid-19 restrictions. We piloted several version of the questions, and then formally tested Covid-19 vs. non Covid-18 framing in our surveys. Conceptually, framing restrictions as related to Covid-19 may lead to subjects justifying these measures as necessary and assigning lower disutility. On the other hand, it may be hard to imagine life with restrictions outside of Covid-19. Our results suggest that very similar responses are obtained with both types of framing. A related concern is that survey respondents may not be able to exactly quantify the relative utility of life with restrictions. We believe that the magnitudes reported here – about a quarter of life quality lost due to light restrictions, and about a third due to severe restrictions – is reasonable. Both states are clearly preferred to paraplegia as a more severe health state by respondents as one may expect. The average utility weight of 0.49 for paraplegia seems well aligned with estimates reported in the literature.^20^ The fourth limitation is that the COVID-19 mortality data available and used to compute the YLL may underestimate the true toll of the epidemic in some countries, particularly in those with limited resources.^21^ However, relative to the global QALY burden reported in this paper, the magnitude of these unaccounted deaths is likely small. Last, we were not able to collect any data on children. Given the absence of a clear age gradient in the valuation of restrictions, applying the same average utility to children seems reasonable. It certainly appears possible that children are disproportionally affected by restrictions on schooling and leisure – future work can hopefully address this question directly.

In summary, the results presented here highlight the very high societal cost of non-pharmaceutical interventions to prevent the spread of infectious diseases such as Covid-19 in terms of quality of life lost. Future policy decisions should take these societal costs into consideration, and try to balance likely reductions in disease transmission from specific measures against their impact on individual and aggregate quality of life.

## Methods

### Study design

This study uses data from cross-sectional surveys conducted in France, India, Italy, the UK and the US to estimate the relative utility of life with and without restrictions, and then computes the national, regional, and global burden of Covid-19 restrictions to date.

To compare this burden to the mortality impact to date, we extracted mortality data from the *Worldometers* website (https://www.worldometers.info/coronavirus/). Data on population size and age structure was taken from the United Nations’ Population Division (https://population.un.org/wpp/). Crude mortality rates by country were taken from the World Development Indicators database (https://data.worldbank.org/). Data on Covid-19 restrictions were retrieved from the Oxford Covid-19 Government Response Tracker (https://doi.org/10.1038/s41562-021-01079-8).^2^

### Survey Participants

Anonymous online surveys were conducted in France, India, Italy, UK and the United State of America using the Amazon Mechanical Turk (MTurk) platform. MTurk is an online platform where volunteer workers sign up for survey or other computational tasks. MTurk has been used in a growing number of studies, and is considered an affordable and reliable source of human participants ^22^. All surveys were completed between June and August 2021.

### Inclusion / Exclusion criteria

All MTurk workers aged 18 and older residents in one of the target countries were invited to participate in the survey. Following MTurk guidelines, survey participants received a small compensation of USD 2 for completing the survey.

### Primary Outcome variables

The primary outcome variable of interest was the total number of quality-adjusted life years lost due to Covid-19 restrictions. Following standard QALY procedures ^19^, we estimated the utility weight associated with each given state through a series of standardized time tradeoff questions (TTOs). In most existing QALY surveys, evaluated states are designed over a specific health condition such as blindness or paraplegia, and survey respondents are then asked to indicate how much utility they get from life with this condition relative to a fully healthy life. Utility weights are then normalized such that perfect health equates to a value of 1, and death is assigned a utility of 0.

In our survey, each state was defined over a set of restrictions. Specifically, we considered the following six restrictions: wearing masks in public spaces; closure of bars, clubs and restaurants; restrictions on international travels; home office; school closures; restrictions on private meetings. In a first step, subjects were asked to complete a series of standard questions related to paraplegia. Paraplegia questions are commonly used in QALY validation studies, and were introduced both to familiarize subjects with time. trade off questions, and to be able to compare average utility weights in this population to those seen in other studies. Next, study participants were introduced to a light and a severe Covid-19 restrictions scenario. Light restrictions included wearing masks in public spaces, restricted access to bars and restaurants and limited international travels. Severe restrictions included all of the light restrictions as well as mandatory home office, remote schooling and the inability to hold private meetings.

We considered two alternative framings for the TTO questions: i) a Covid-19 specific framing, in which we asked subjects to trade off 12 months under a specified set of restrictions against *x* months of their usual life (with *x* ranging between 0 and 12); ii) a more neutral framing, in which we asked subjects to trade off x years of healthy life against 10 years of life with specific restrictions. No differences were found between neutral (end-of-life) and Covid-19 specific framing (Appendix Figure A1).

The original survey questions (in English) are provided in Supplemental Materials Table 2. Translations to French and Italian were made by the research team.

In order to quantify the relative magnitude of specific measures as well as respondents’ willingness to pay (WTP) to avoid specific restrictions, we implemented a discrete choice experiment (DCE), in which we asked subjects to choose between bundles of living conditions involving restrictions on everyday life as well as pre-specified incomes. Full details on the specific questions asked as well as the choice sets given to study participants are provided in the Supplemental Materials.

### Statistical Analysis

We started by estimating average utility weights with mild and severe restrictions in the pooled sample, and compared it to the reported utility weights for paraplegia. We estimated utility weights for the sample overall, as well as by country, gender and age group.

Then, we combined our utility estimates with data on population size and data on the duration of light and severe Covid-19 restrictions in each country to generate national, regional and global estimates of the total utility loss to date. We used the latest mortality estimates s from Worldometers to quantify the total Years of Life Lost (YLL) from Covid-19 to date (accessed on 6^th^ of September 2021). To calculate the YLL due to Covid-19 for all countries, we took conditional life expectancies from Pifarré et al ^1^ and multiplied these average life expectancies with the reported Covid-19 deaths to date. For countries where no data on conditional life expectancy was available, we used the mean life expectancies reported in the sub-region. No data was available for any central Asian country and Melanesia. We thus used average western Asian life expectancy for central Asia, and used estimates from Oceania for Melanesia. To account for pre-existing morbidities in the general population, we used average QALY estimates from Love-Koh, et al. ^23^ as reference.

To estimate the relative disutility from each specific measure, we analyzed responses from the DCE using standard logistic regression models. Estimated marginal effects in the choice model were scaled by the marginal effects obtained for the median income to obtain estimated WTP for (preventing) restrictions.

### Ethical Considerations

All surveys were completed anonymously online. All respondents provided consent to the use of data for research by ticking a box before the questionnaire starts. Due to the absence of identifiable data, the study was rated as non-human subjects research by the ethics commission (EKNZ Req 2021.00616).

## Data Availability

All data will be made available by the authors.

## Data availability

All data will be made available by the authors.

## Code availability

All code necessary to reproduce this analysis will be uploaded to a public repository.

