## Supplemental Materials for "Global Burden of Covid-19 Restrictions: National, Regional and Global Estimates"

**Supplemental Materials Figure S1: Mean Utility Weight by Framing**

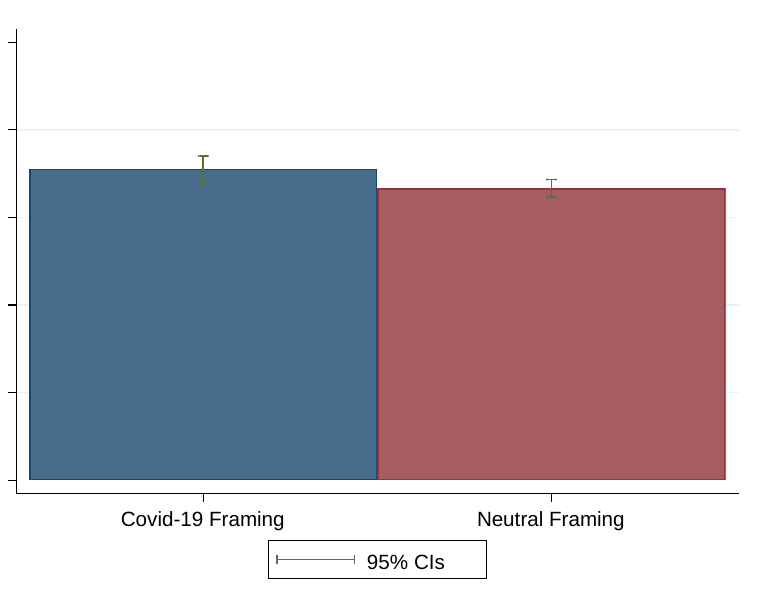

**Supplemental Materials Figure S2: Distribution of Individual Utility Weights overall and by Country**

| 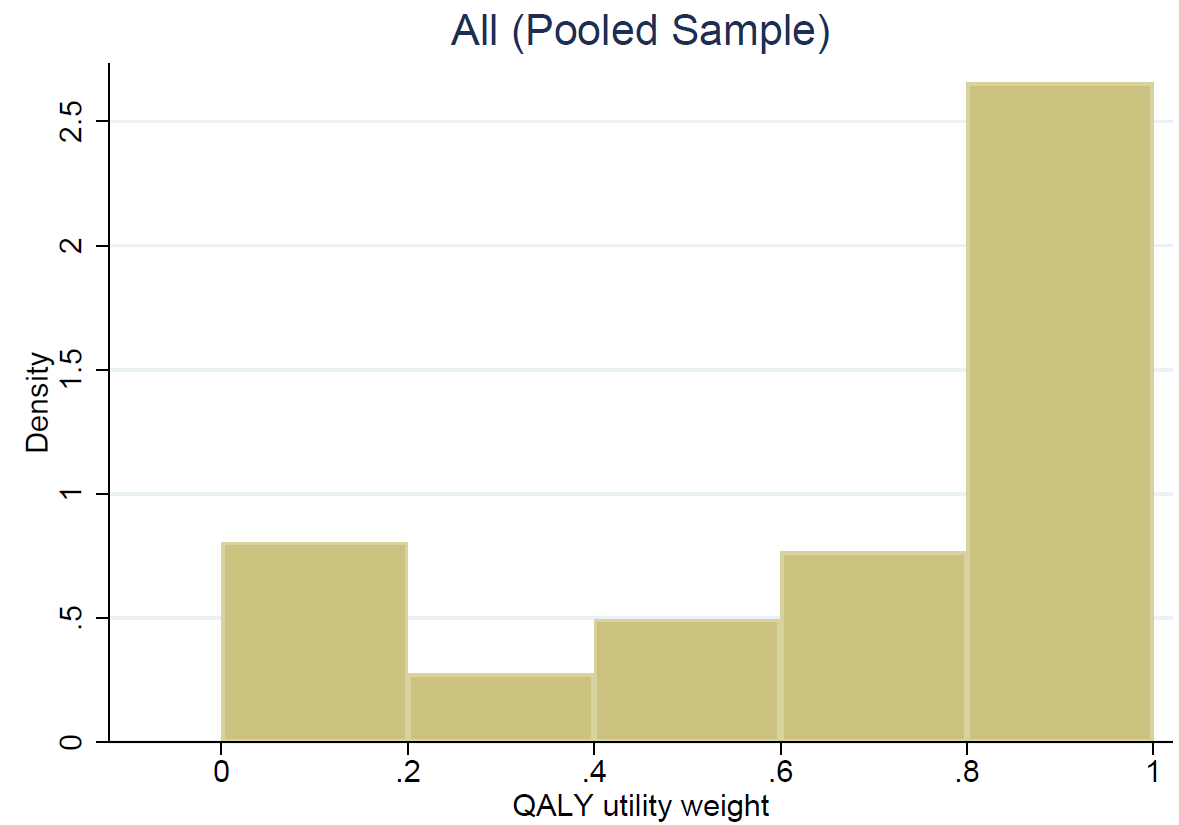 | 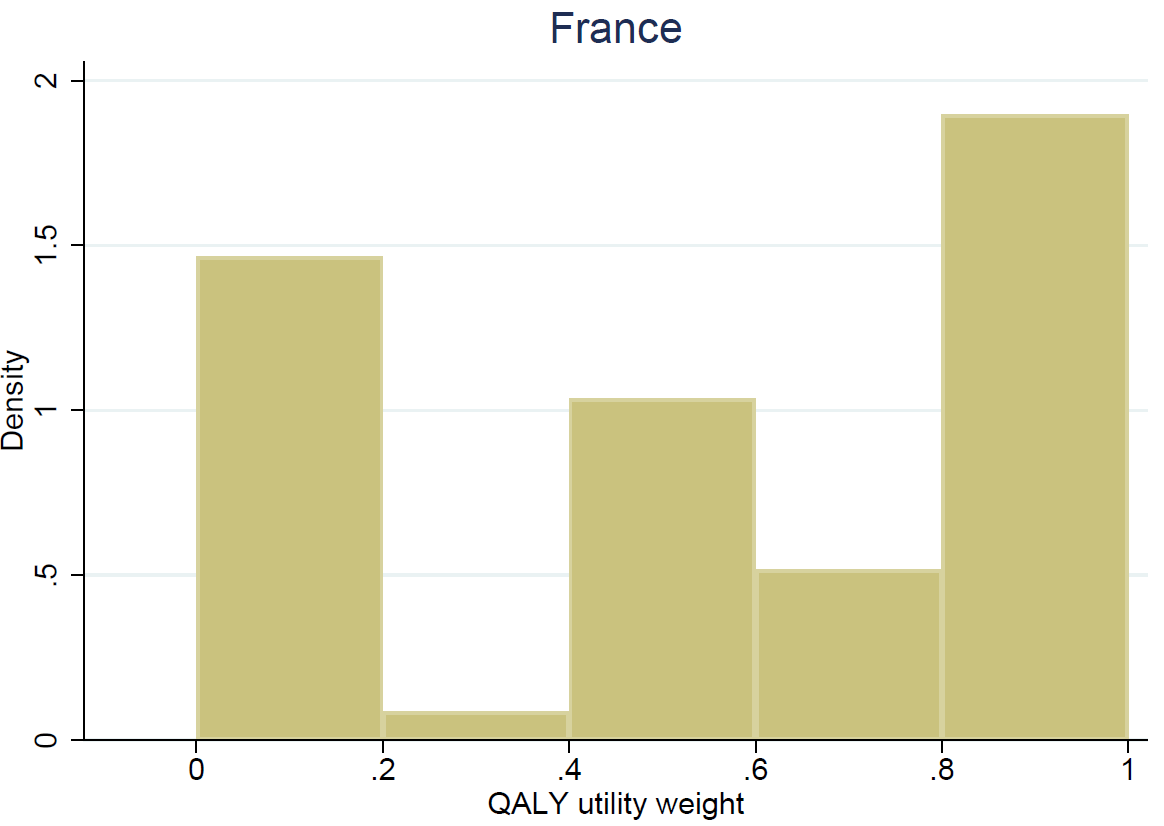 |
| --- | --- |
| 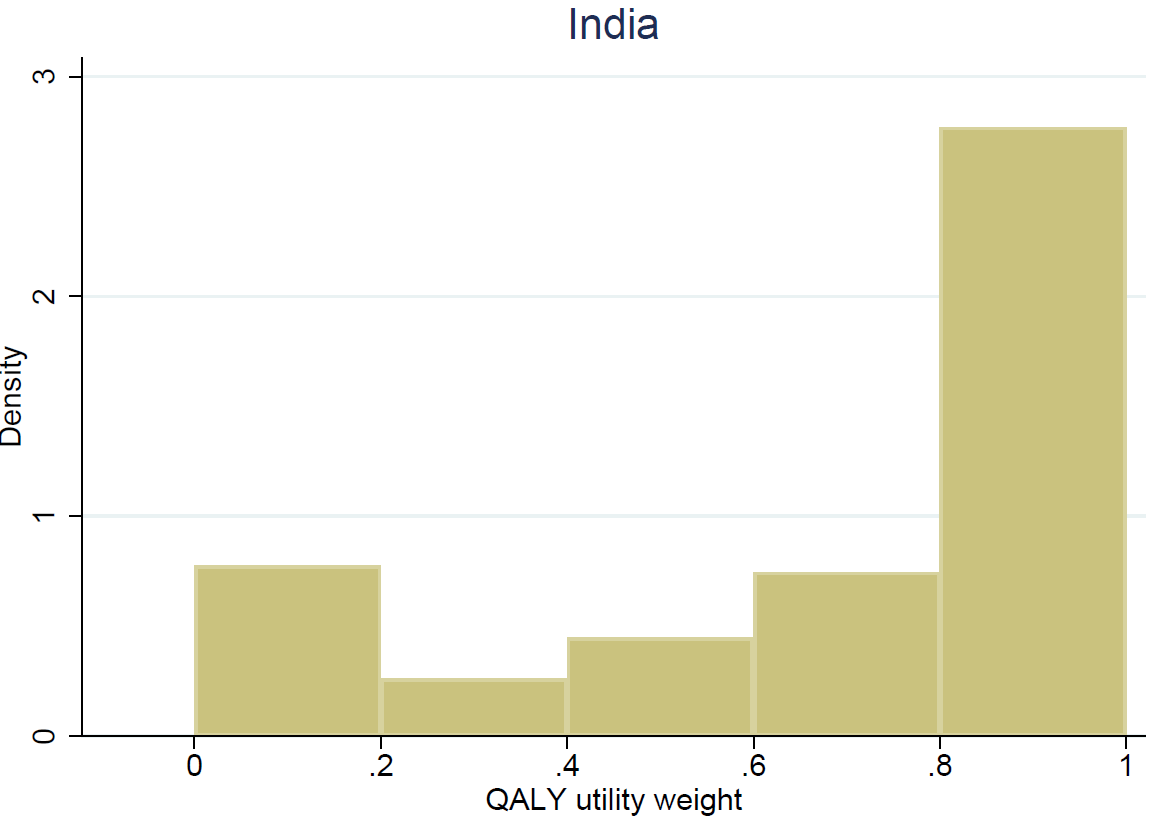 | 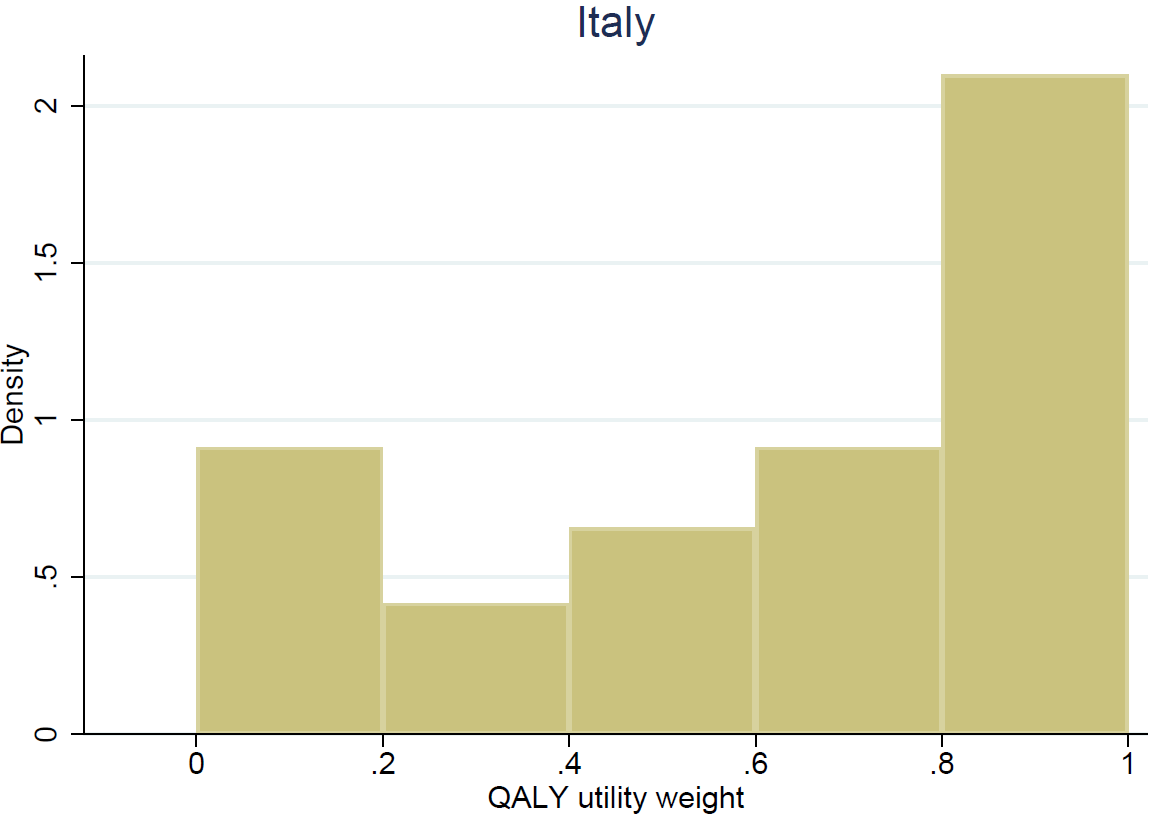 |
| 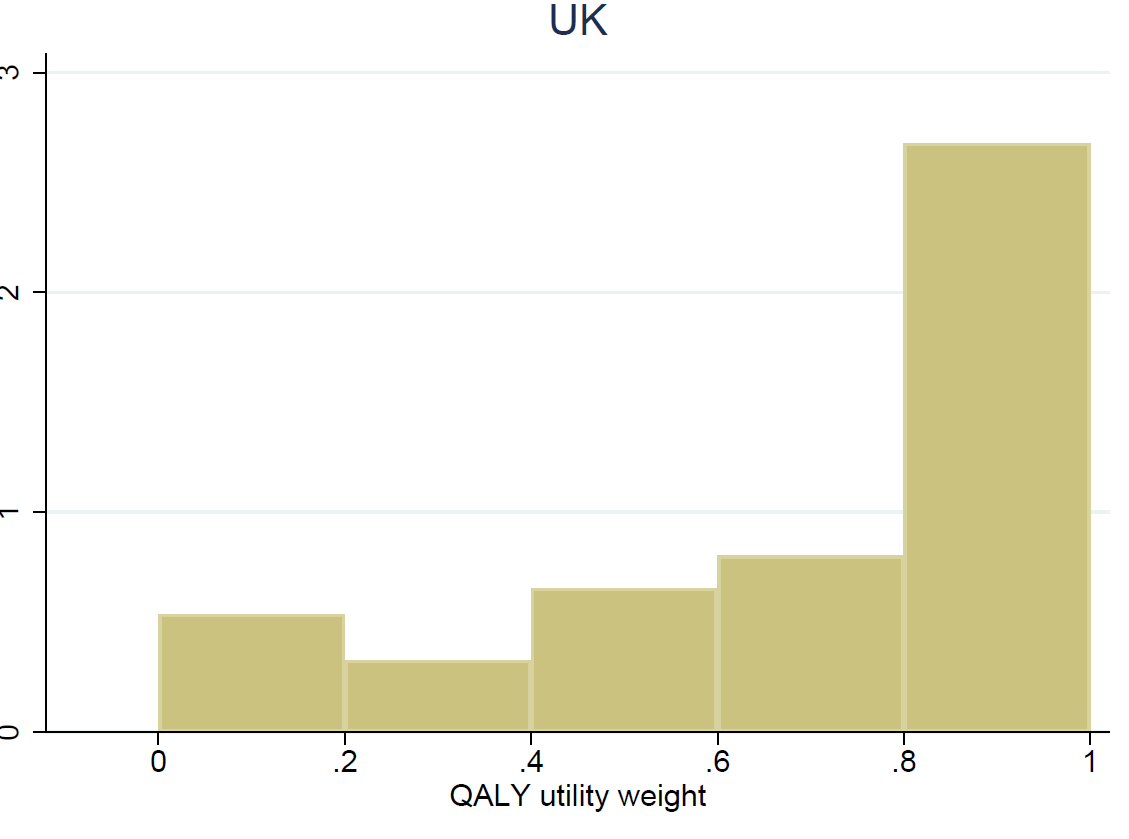 | 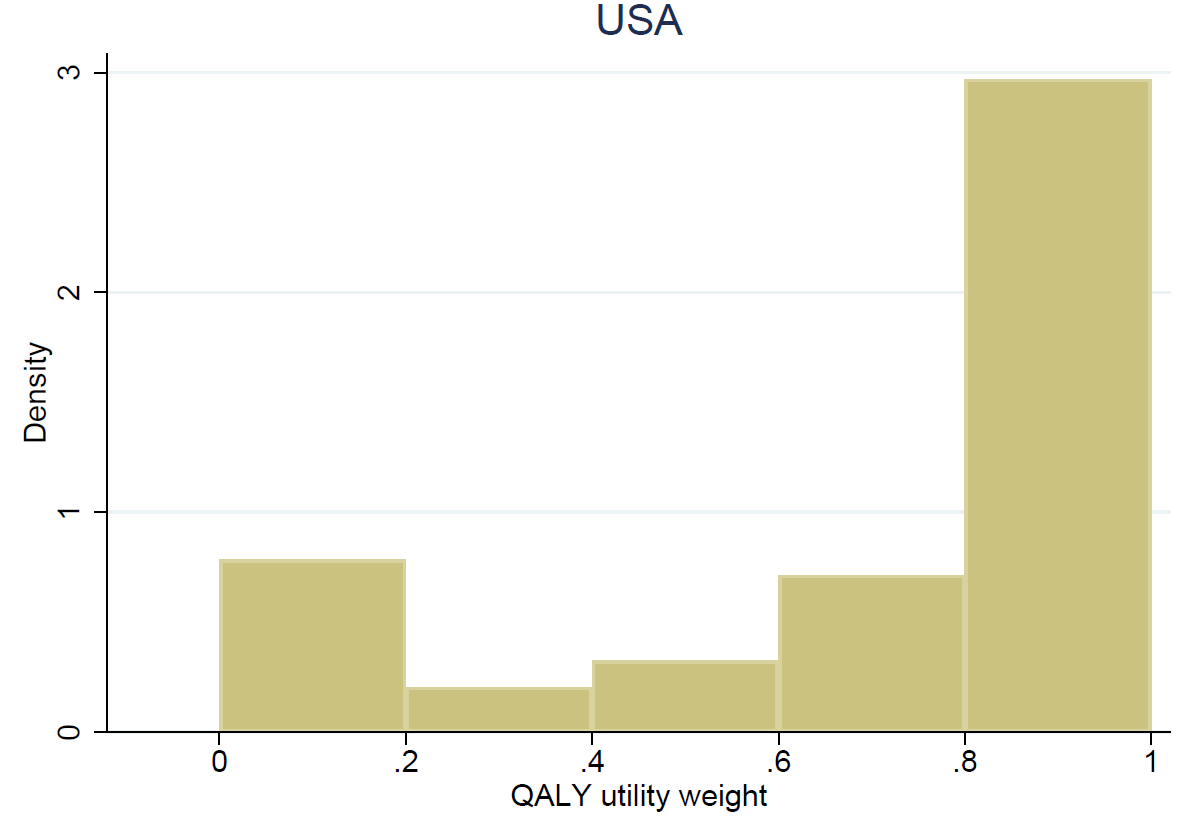 |

**Supplemental Materials Figure S3: Months of Light Restrictions**

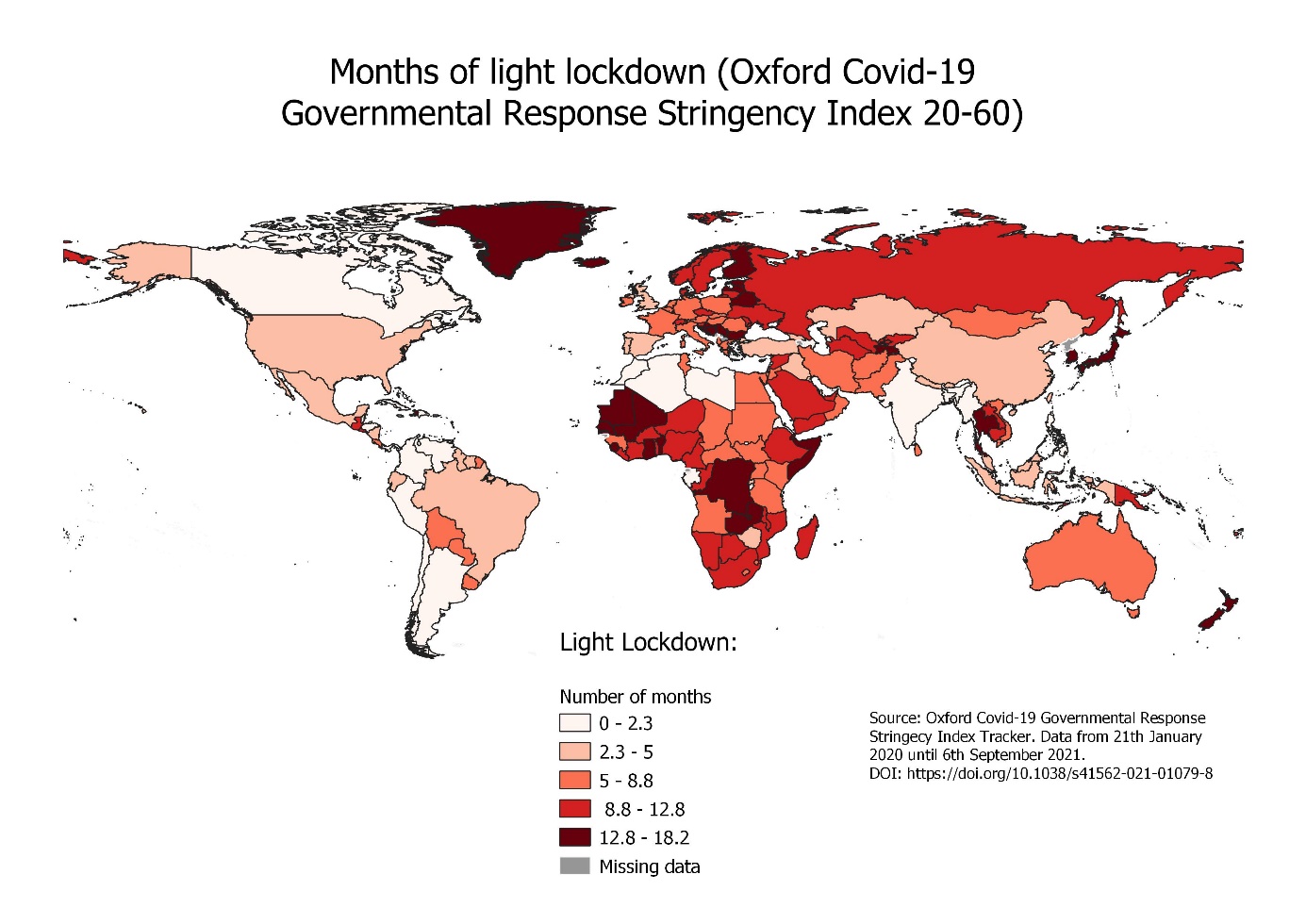

**Supplemental Materials Figure S4: Months of Severe Restrictions**

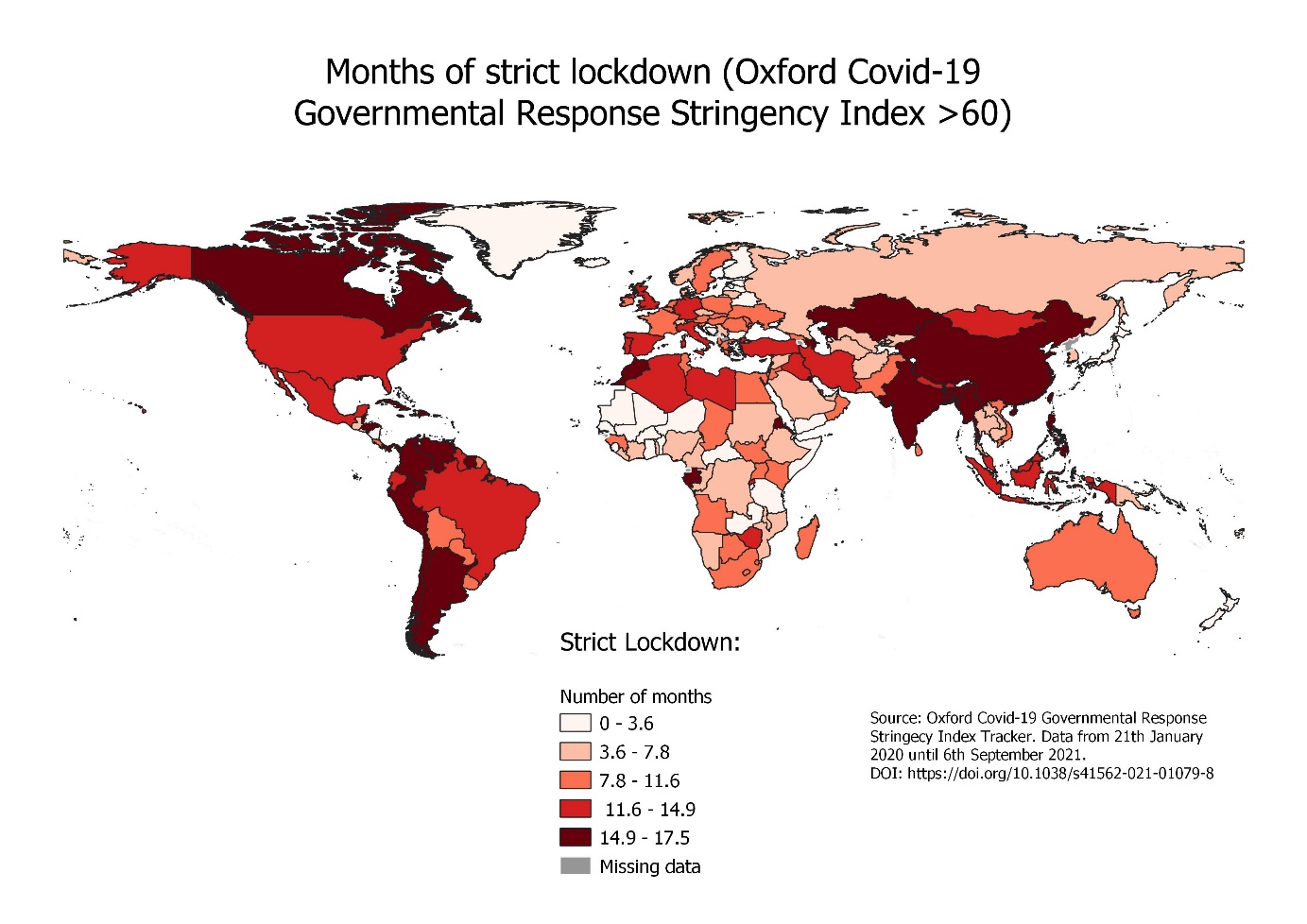

**Supplemental Material Table S1: Characteristics of Survey Participants**

| Country | USA | India | UK | Italy | France |
| --- | --- | --- | --- | --- | --- |
| Total respondents | 298 | 228 | 192 | 181 | 33 |
| Female | 100 | 89 | 77 | 57 | 8 |
| Male | 198 | 139 | 115 | 124 | 25 |
| Age group: <20 | 0 | 0 | 5 | 9 | 0 |
| Age group: 20-29 | 53 | 78 | 58 | 93 | 8 |
| Age group: 30-39 | 110 | 98 | 83 | 35 | 16 |
| Age group: 40-49 | 56 | 24 | 32 | 24 | 5 |
| Age group: 50-59 | 39 | 18 | 11 | 24 | 4 |
| Age group: 60-69 | 33 | 7 | 2 | 18 | 0 |
| Age group: 70+ | 7 | 3 | 1 | 2 | 0 |
| Nationality: citizen | 297 | 214 | 166 | 167 | 30 |
| Nationality: foreign | 1 | 14 | 26 | 14 | 3 |
| Highest level of educ: Upper secondary or lower | 35 | 31 | 42 | 69 | 0 |
| Highest level of educ: Post secondary | 208 | 152 | 99 | 102 | 2 |
| Highest level of educ: Tertiary | 55 | 45 | 51 | 10 | 30 |
| Children under age 6: No children | 192 | 98 | 146 | 168 | 29 |
| Children under age 6: One child | 87 | 85 | 37 | 10 | 3 |
| Children under age 6:Two children | 15 | 33 | 8 | 2 | 1 |
| Children under age 6: More than two children | 4 | 12 | 1 | 1 | 0 |
| Children age 6-17: No children | 192 | 88 | 147 | 167 | 27 |
| Children age 6-17: One child | 63 | 97 | 24 | 8 | 3 |
| Children age 6-17: Two children | 34 | 29 | 20 | 5 | 2 |
| Children age 6-17: More than two children | 9 | 14 | 1 | 1 | 1 |
| Income group: Lowest | 55 | 29 | 23 | 71 | 6 |
| Income group: Lower | 94 | 62 | 31 | 28 | 16 |
| Income group: Medium | 78 | 29 | 51 | 33 | 5 |
| Income group: Higher | 48 | 30 | 43 | 16 | 1 |
| Income group: Highest | 23 | 76 | 36 | 8 | 3 |
| Income group: Refused | 0 | 2 | 8 | 25 | 2 |
| Job loss last 12m: Yes | 68 | 112 | 27 | 21 | 6 |
| Job loss last 12m: No | 227 | 113 | 164 | 158 | 27 |
| Covid infection: Yes | 55 | 78 | 24 | 14 | 6 |
| Covid infection: No | 243 | 150 | 168 | 167 | 27 |

**Supplemental Material Table S2: Survey Questions**

***Time-Tradeoff Questions: Covid Framing***

*Light Restrictions*

*“Now please consider instead another restrictions scenario, where* ***you have to wear a mask at all times in public, you are not allowed to go out for dining, drinks, clubs or to the gym, and travelling is prohibited.*** *If you had to make a choice between life with these restrictions and your usual life: “*

*Severe Restrictions*

*Now please consider instead an even stricter lockdown scenario, where* ***you have to wear masks in all public spaces, you cannot go out for dining, drinks, clubs or to the gym, private parties and events are prohibited, all children have to be taught remotely while staying at home, and you are not allowed to travel.*** *If you had to make a choice between life in this type of lockdown and your usual life:*

***Time-Tradeoff Questions: Neutral End of Life Framing***

*“We would now like to ask you to make (hypothetical) choices between life under different circumstances. Specifically, we will ask you to choose between a certain quantity of time left to live with restrictions and a (typically shorter) quantity of time left to live without restrictions. Assume that all other things are the same, i.e., consider only your private situation.”*

*Assume a world without COVID-19, and that you have only a limited amount of time left to live. You have to make a decision between living in one of the following two countries: Country A with no restrictions, and country B, where private life is restricted.* ***Specifically, country B requires you to wear a mask at all times in public, does not allow you to go out for dining, drinks, clubs or to the gym, and does not allow traveling****.*

..would you rather have **[X]** years of your usual life (Option A) or **10** years of life with restrictions in country B (Option B)?” (with X ranging from 0-10)

Following standard QALY procedures, we sequentially lowered the number of healthy years [X] offered until subjects expressed they were either indifferent or preferred life with restrictions. To quantify indifference points for subjects switching between cutoffs, we used linear interpolation, such that subjects preferring 6 years of their usual life to 10 years of restricted life, but preferring 10 years of restricted life to 4 years of usual life were coded at the interval midpoint 5.

**Discrete Choice Experiment**

In order to assess the relative burden of each individual restriction, we also developed a discrete choice experiment, where subjects had to choose between specific bundles of restrictions. We considered six main restrictions: 1) having to wear a mask in all public spaces, including offices 2) not being able to hold or attend events (e.g. private parties, concerts, weddings)? 3) not being able to go to the gym or fitness classes 4) not being able to go out to restaurants, bars and clubs 5) not being able to travel and 6) not being able to send kids to daycare or school (when these were closed)? In order to be able to directly quantify the relative burden of each restriction, we also added a continuous wage variable to the choice sets, and asked subjects to choose between two hypothetical places to live as illustrated in the examples shown in Figure 1:

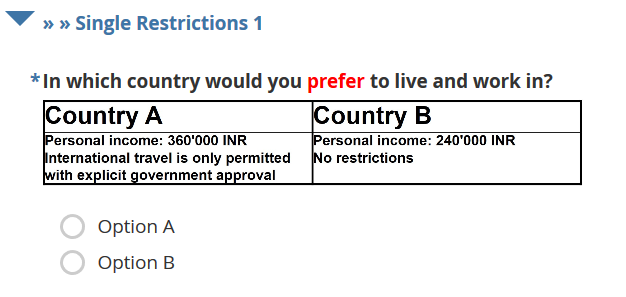

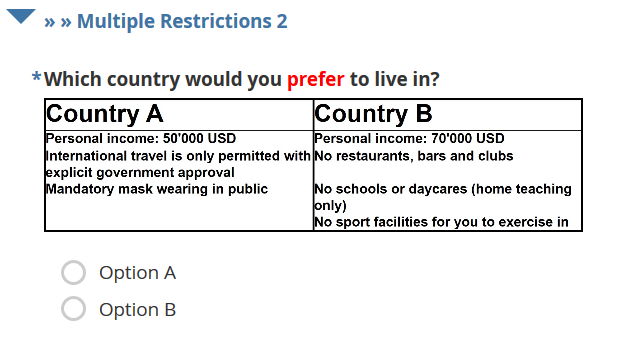

Figure 1 legend: Example of a single restriction DCE from the survey in India (topt) and a multiple restriction DCE from the survey in the US (bottom)

**Supplemental Material Table S3: Country-level Restriction Exposure, life years and QALYs lost between January 2020 and August 2021**

| Country | Population (Millions) | Months of light restrictions^a)b)^ | Months of severe restrictions^a)b)^ | Estimated QALY loss due to restrictions (Millions) | 95% confidence interval (Millions) | Life years lost due to Covid-19 mortality (Millions) | Ratio QALYs to life years lost due to Covid-19 mortality |
| --- | --- | --- | --- | --- | --- | --- | --- |
| China | 1439.32 | 2.8 | 14.4 | 6081.36 | 5645.06 - 6517.66 | 0.65 | 9356 |
| India | 1393.75 | 1.3 | 14.5 | 5619.47 | 5219.63 - 6019.31 | 98.46 | 57 |
| United States | 332.97 | 3 | 12.3 | 1238.26 | 1149.11 - 1327.41 | 82.3 | 15 |
| Indonesia | 276.45 | 4.9 | 11.9 | 1136.53 | 1053.88 - 1219.19 | 31.43 | 36 |
| Pakistan | 225.23 | 7.1 | 8.7 | 857.14 | 793.71 - 920.57 | 5.86 | 146 |
| Brazil | 214.09 | 1.8 | 13.5 | 820.73 | 762.12 - 879.34 | 114.91 | 7 |
| Nigeria | 211.3 | 9.5 | 5.9 | 764.23 | 706.58 - 821.89 | 0.46 | 1661 |
| Bangladesh | 166.35 | 0.1 | 15.2 | 660.64 | 613.99 - 707.29 | 5.95 | 111 |
| Russia | 146 | 10.7 | 5.2 | 512.19 | 473.29 - 551.09 | 28.04 | 18 |
| Mexico | 130.31 | 4 | 11.2 | 487 | 451.69 - 522.31 | 51.81 | 9 |
| Japan | 126.08 | 16.3 | 0 | 412.25 | 379.62 - 444.88 | 2.28 | 181 |
| Ethiopia | 117.84 | 8.6 | 6.8 | 430.8 | 398.52 - 463.08 | 0.86 | 501 |
| Philippines | 111.06 | 1.7 | 14.8 | 468.3 | 434.91 - 501.69 | 7.91 | 59 |
| Egypt | 104.29 | 3.9 | 10.7 | 378.33 | 350.89 - 405.78 | 3.04 | 124 |
| Vietnam | 98.23 | 8.2 | 8.6 | 386.18 | 357.48 - 414.89 | 3.08 | 125 |
| Democratic Republic of Congo | 92.31 | 10.9 | 3.9 | 313.54 | 289.54 - 337.54 | 0.19 | 1650 |
| Turkey | 85.26 | 2 | 13.7 | 336.13 | 312.1 - 360.16 | 9.68 | 35 |
| Iran | 85.08 | 6.2 | 9.9 | 328.95 | 304.78 - 353.12 | 24.85 | 13 |
| Germany | 84.06 | 5.2 | 10.7 | 311.48 | 288.74 - 334.22 | 9.23 | 34 |
| Thailand | 69.98 | 11.9 | 3.6 | 235.36 | 217.28 - 253.45 | 3 | 78 |
| United Kingdom | 68.25 | 1.3 | 14 | 257.78 | 239.43 - 276.12 | 17.72 | 15 |
| France | 65.42 | 6.1 | 9.8 | 241.57 | 223.82 - 259.31 | 11.42 | 21 |
| Tanzania | 61.45 | 6 | 0 | 79.89 | 73.57 - 86.21 | 0.01 | 7989 |
| Italy | 60.37 | 4.7 | 11.4 | 227.07 | 210.55 - 243.58 | 16.11 | 14 |
| South Africa | 60.07 | 8.1 | 7.1 | 214.45 | 198.42 - 230.47 | 15.01 | 14 |
| Kenya | 54.97 | 6.2 | 8.6 | 198.58 | 183.93 - 213.22 | 0.86 | 231 |
| Myanmar | 54.78 | 0.6 | 15 | 219.48 | 203.93 - 235.02 | 3.66 | 60 |
| Colombia | 51.43 | 0.4 | 14.9 | 201.33 | 187.09 - 215.58 | 24.67 | 8 |
| South Korea | 51.31 | 12.9 | 3.9 | 185.2 | 170.97 - 199.43 | 0.32 | 579 |
| Uganda | 47.2 | 7.7 | 7.5 | 173.28 | 160.38 - 186.19 | 0.55 | 315 |
| Spain | 46.77 | 2.2 | 13.5 | 177.68 | 164.97 - 190.4 | 10.55 | 17 |
| Argentina | 45.62 | 0.3 | 15.3 | 181.74 | 168.89 - 194.58 | 22.15 | 8 |
| Sudan | 44.88 | 7 | 7.3 | 154.17 | 142.71 - 165.63 | 0.51 | 302 |
| Algeria | 44.65 | 0.4 | 14.5 | 171.81 | 159.65 - 183.96 | 0.98 | 175 |
| Ukraine | 43.47 | 7.4 | 8.1 | 153.95 | 142.52 - 165.38 | 8.06 | 19 |
| Iraq | 41.13 | 4.5 | 11.6 | 165.66 | 153.63 - 177.69 | 3.52 | 47 |
| Afghanistan | 39.81 | 6 | 4.7 | 101.31 | 93.72 - 108.91 | 1.6 | 63 |
| Canada | 38.08 | 0.2 | 15.4 | 148.57 | 138.08 - 159.07 | 3.34 | 44 |
| Poland | 37.8 | 6.4 | 9.2 | 136.42 | 126.37 - 146.48 | 11.24 | 12 |
| Morocco | 37.35 | 0.2 | 15.1 | 147.19 | 136.79 - 157.59 | 2.36 | 62 |
| Saudi Arabia | 35.36 | 9.7 | 6 | 127.91 | 118.26 - 137.56 | 1.43 | 89 |
| Uzbekistan | 33.96 | 10.2 | 5 | 124.99 | 115.89 - 134.09 | 0.23 | 543 |
| Angola | 33.9 | 4.5 | 10.2 | 117.85 | 108.9 - 126.8 | 0.19 | 620 |
| Peru | 33.44 | 0.5 | 15.1 | 133.58 | 124.13 - 143.04 | 39.08 | 3 |
| Malaysia | 32.79 | 4.4 | 11.4 | 127.1 | 117.87 - 136.33 | 4.26 | 30 |
| Mozambique | 32.14 | 8.4 | 6.9 | 117.35 | 108.57 - 126.13 | 0.34 | 345 |
| Ghana | 31.73 | 13.5 | 1.9 | 108.03 | 99.61 - 116.44 | 0.19 | 569 |
| Yemen | 30.49 | 13.9 | 0 | 91.63 | 84.38 - 98.88 | 0.25 | 367 |
| Nepal | 29.67 | 3.9 | 11.8 | 116.07 | 107.67 - 124.47 | 2.43 | 48 |
| Madagascar | 28.41 | 6.8 | 8.3 | 104.02 | 96.33 - 111.72 | 0.17 | 612 |
| Venezuela | 28.35 | 0.1 | 14.8 | 108.58 | 100.91 - 116.25 | 0.81 | 134 |
| Cameroon | 27.21 | 10.6 | 4.8 | 97.02 | 89.64 - 104.41 | 0.24 | 404 |
| Cote d'Ivoire | 27.04 | 8.5 | 4.2 | 79.77 | 73.71 - 85.82 | 0.08 | 997 |
| Australia | 25.8 | 6.3 | 9.1 | 92.87 | 86.03 - 99.72 | 0.11 | 844 |
| Niger | 25.09 | 10.5 | 1.5 | 67.34 | 62.1 - 72.59 | 0.04 | 1684 |
| Sri Lanka | 21.5 | 7.2 | 7.8 | 60.74 | 56.05 - 65.43 | 0.03 | 2025 |
| Burkina Faso | 21.48 | 10.1 | 2.4 | 75.13 | 69.55 - 80.71 | 2.26 | 33 |
| Mali | 20.84 | 13.8 | 1.5 | 70.77 | 65.24 - 76.3 | 0.1 | 708 |
| Malawi | 19.63 | 10.6 | 4.5 | 68.54 | 63.32 - 73.77 | 0.4 | 171 |
| Chile | 19.28 | 0.3 | 14.7 | 73.32 | 68.14 - 78.51 | 7.3 | 10 |
| Romania | 19.11 | 6.9 | 8.6 | 67.91 | 62.89 - 72.94 | 5.18 | 13 |
| Kazakhstan | 19 | 1.5 | 14 | 74.56 | 69.25 - 79.88 | 1.64 | 45 |
| Zambia | 18.9 | 14.2 | 0.9 | 62.82 | 57.89 - 67.75 | 0.65 | 97 |
| Guatemala | 18.25 | 9.3 | 6.5 | 67.52 | 62.45 - 72.6 | 2.41 | 28 |
| Syria | 17.93 | 10 | 5.5 | 64.14 | 59.28 - 68.99 | 0.34 | 189 |
| Ecuador | 17.91 | 4.1 | 11.3 | 67.99 | 63.06 - 72.92 | 6.37 | 11 |
| Senegal | 17.19 | 11.6 | 3.1 | 57.24 | 52.83 - 61.65 | 0.33 | 173 |
| Netherlands | 17.17 | 4.4 | 11.1 | 62.99 | 58.42 - 67.57 | 1.79 | 35 |
| Cambodia | 16.95 | 11.8 | 3.7 | 59.05 | 54.52 - 63.59 | 0.45 | 131 |
| Chad | 16.9 | 4.8 | 9.7 | 61.21 | 56.73 - 65.68 | 0.03 | 2040 |
| Somalia | 16.34 | 11.9 | 1.8 | 50.01 | 46.11 - 53.9 | 0.18 | 278 |
| Zimbabwe | 15.08 | 3.5 | 11.7 | 58.01 | 53.82 - 62.2 | 0.81 | 72 |
| Guinea | 13.49 | 7.2 | 7.8 | 48.96 | 45.32 - 52.59 | 0.06 | 816 |
| Rwanda | 13.28 | 1.8 | 13.9 | 53.87 | 50.02 - 57.71 | 0.2 | 269 |
| Benin | 12.44 | 13.9 | 1.4 | 41.99 | 38.71 - 45.28 | 0.02 | 2100 |
| Burundi | 12.25 | 1 | 0 | 2.66 | 2.45 - 2.87 | 0.01 | 266 |
| Tunisia | 11.94 | 6.2 | 9.3 | 44.04 | 40.8 - 47.28 | 4.31 | 10 |
| Bolivia | 11.83 | 4.8 | 8.5 | 38.22 | 35.42 - 41.02 | 3.64 | 11 |
| Belgium | 11.64 | 6.5 | 9 | 41.67 | 38.6 - 44.74 | 2.52 | 17 |
| Haiti | 11.54 | 11.3 | 4 | 39.95 | 36.9 - 43.01 | 0.12 | 333 |
| South Sudan | 11.33 | 5.6 | 9.8 | 42.95 | 39.81 - 46.1 | 0.02 | 2148 |
| Cuba | 11.32 | 2.1 | 13 | 41.76 | 38.77 - 44.74 | 1.12 | 37 |
| Dominican Republic | 10.96 | 0.1 | 15.4 | 43.89 | 40.79 - 46.99 | 0.79 | 56 |
| Czechia | 10.73 | 8.5 | 7.1 | 37.62 | 34.81 - 40.44 | 4.54 | 8 |
| Greece | 10.37 | 5.4 | 10.2 | 37.5 | 34.76 - 40.25 | 1.73 | 22 |
| Jordan | 10.31 | 3.8 | 10.8 | 37.51 | 34.79 - 40.22 | 1.74 | 22 |
| Azerbaijan | 10.23 | 0.5 | 15.4 | 41.59 | 38.64 - 44.53 | 0.98 | 42 |
| Portugal | 10.17 | 1.8 | 13.7 | 36.44 | 33.74 - 39.14 | 1.95 | 19 |
| Sweden | 10.16 | 7 | 8.6 | 38.14 | 35.42 - 40.86 | 2.21 | 17 |
| Honduras | 10.06 | 0.1 | 15.4 | 40.76 | 37.88 - 43.64 | 1.78 | 23 |
| United Arab Emirates | 10.01 | 11.4 | 4.3 | 35.51 | 32.79 - 38.22 | 0.34 | 104 |
| Tajikistan | 9.75 | 14.2 | 0.4 | 30.8 | 28.37 - 33.23 | 0.02 | 1540 |
| Hungary | 9.64 | 6.3 | 9.3 | 34.72 | 32.16 - 37.27 | 4.49 | 8 |
| Belarus | 9.45 | 13.6 | 0 | 26.36 | 24.28 - 28.45 | 0.57 | 46 |
| Israel | 9.33 | 5 | 10.8 | 35.54 | 32.95 - 38.13 | 1.2 | 30 |
| Papua New Guinea | 9.12 | 10.4 | 4.7 | 31.61 | 29.21 - 34.02 | 0.02 | 1581 |
| Austria | 9.06 | 5.6 | 10.1 | 33.15 | 30.72 - 35.58 | 1.07 | 31 |
| Switzerland | 8.72 | 9.8 | 6 | 30.5 | 28.2 - 32.81 | 1.09 | 28 |
| Serbia | 8.7 | 11.5 | 4.1 | 29.41 | 27.16 - 31.67 | 0.92 | 32 |
| Togo | 8.47 | 10 | 5.2 | 29.95 | 27.68 - 32.22 | 0.04 | 749 |
| Sierra Leone | 8.14 | 12.5 | 2.6 | 27.58 | 25.44 - 29.71 | 0.02 | 1379 |
| Laos | 7.38 | 9.4 | 3.6 | 21.81 | 20.14 - 23.48 | 0 | #DIV/0! |
| Paraguay | 7.22 | 5.7 | 9.4 | 26.41 | 24.47 - 28.35 | 3.13 | 8 |
| Libya | 6.97 | 1.5 | 13.9 | 27.4 | 25.45 - 29.35 | 0.79 | 35 |
| Bulgaria | 6.9 | 12.5 | 3.2 | 22.97 | 21.2 - 24.74 | 2.85 | 8 |
| Lebanon | 6.79 | 2.9 | 12.9 | 26.72 | 24.8 - 28.64 | 1.35 | 20 |
| Nicaragua | 6.7 | 2.5 | 0 | 3.57 | 3.29 - 3.85 | 0.04 | 89 |
| Kyrgyzstan | 6.63 | 8.8 | 7 | 24.56 | 22.72 - 26.39 | 0.43 | 57 |
| El Salvador | 6.52 | 9.2 | 6.4 | 23.45 | 21.69 - 25.21 | 0.58 | 40 |
| Singapore | 5.9 | 14.8 | 2.4 | 21.5 | 19.83 - 23.17 | 0.01 | 2150 |
| Denmark | 5.81 | 9 | 6.9 | 20.73 | 19.17 - 22.28 | 0.34 | 61 |
| Congo | 5.66 | 9 | 6 | 19.95 | 18.45 - 21.45 | 0.03 | 665 |
| Finland | 5.55 | 13.1 | 2.4 | 18.12 | 16.71 - 19.52 | 0.14 | 129 |
| Norway | 5.46 | 8.9 | 6.6 | 19.07 | 17.64 - 20.51 | 0.11 | 173 |
| Slovakia | 5.46 | 6.8 | 8.9 | 19.81 | 18.35 - 21.28 | 1.87 | 11 |
| Oman | 5.24 | 4.9 | 10.6 | 20.04 | 18.58 - 21.5 | 0.68 | 29 |
| Liberia | 5.18 | 8.6 | 6.8 | 18.9 | 17.48 - 20.32 | 0.03 | 630 |
| Costa Rica | 5.14 | 6 | 9.6 | 19.08 | 17.67 - 20.48 | 1.1 | 17 |
| New Zealand | 5 | 12.6 | 2.7 | 18.67 | 17.31 - 20.03 | 0.68 | 27 |
| Ireland | 4.99 | 4.6 | 11 | 16.39 | 15.12 - 17.66 | 0 | #DIV/0! |
| Central African Republic | 4.91 | 7.1 | 3.6 | 12.24 | 11.31 - 13.17 | 0.02 | 612 |
| Mauritania | 4.77 | 11.5 | 3.5 | 16.25 | 15.01 - 17.5 | 0.13 | 125 |
| Panama | 4.38 | 1.8 | 13.3 | 16.7 | 15.51 - 17.9 | 1.4 | 12 |
| Kuwait | 4.33 | 1.7 | 14.2 | 17.39 | 16.15 - 18.63 | 0.4 | 43 |
| Croatia | 4.08 | 12.8 | 2.8 | 13.44 | 12.4 - 14.48 | 1.04 | 13 |
| Moldova | 4.02 | 8.1 | 7.6 | 14.48 | 13.4 - 15.56 | 0.96 | 15 |
| Georgia | 3.98 | 4.3 | 11.4 | 14.97 | 13.88 - 16.06 | 1.31 | 11 |
| Eritrea | 3.6 | 0.4 | 15.1 | 14.61 | 13.57 - 15.64 | 0.01 | 1461 |
| Uruguay | 3.49 | 6.7 | 8.8 | 12.61 | 11.67 - 13.54 | 1.19 | 11 |
| Mongolia | 3.33 | 4.3 | 12.8 | 14.15 | 13.12 - 15.17 | 0.13 | 109 |
| Bosnia and Herzegovina | 3.26 | 13 | 2.5 | 10.68 | 9.85 - 11.51 | 1.23 | 9 |
| Jamaica | 2.97 | 0.3 | 15 | 11.65 | 10.82 - 12.47 | 0.32 | 36 |
| Albania | 2.87 | 6 | 9.8 | 10.68 | 9.89 - 11.46 | 0.31 | 34 |
| Qatar | 2.81 | 1.9 | 13.5 | 10.98 | 10.19 - 11.76 | 0.1 | 110 |
| Lithuania | 2.68 | 8.8 | 6.5 | 9.1 | 8.42 - 9.79 | 0.61 | 15 |
| Namibia | 2.59 | 10 | 5.2 | 9.11 | 8.42 - 9.8 | 0.61 | 15 |
| Gambia | 2.49 | 8.6 | 6.6 | 8.99 | 8.31 - 9.66 | 0.06 | 150 |
| Botswana | 2.4 | 8 | 7 | 8.52 | 7.88 - 9.15 | 0.41 | 21 |
| Gabon | 2.28 | 0.2 | 15.3 | 9.27 | 8.62 - 9.93 | 0.03 | 309 |
| Lesotho | 2.16 | 6 | 8.9 | 7.79 | 7.22 - 8.37 | 0.07 | 111 |
| Slovenia | 2.08 | 7.3 | 8.1 | 7.27 | 6.73 - 7.81 | 0.55 | 13 |
| Latvia | 1.86 | 12.2 | 3.4 | 6.19 | 5.71 - 6.67 | 0.34 | 18 |
| Bahrain | 1.76 | 7.5 | 8.4 | 6.68 | 6.18 - 7.17 | 0.23 | 29 |
| Trinidad and Tobago | 1.4 | 0.3 | 14.8 | 5.37 | 4.99 - 5.75 | 0.26 | 21 |
| Timor | 1.34 | 9.5 | 5.7 | 4.76 | 4.4 - 5.12 | 0.02 | 238 |
| Estonia | 1.33 | 12.4 | 3.2 | 4.41 | 4.07 - 4.75 | 0.17 | 26 |
| Mauritius | 1.27 | 2.2 | 6.3 | 2.61 | 2.43 - 2.8 | 0.01 | 261 |
| Cyprus | 1.22 | 5.3 | 10.1 | 4.44 | 4.12 - 4.77 | 0.09 | 49 |
| Eswatini | 1.17 | 5.5 | 9.9 | 4.44 | 4.12 - 4.77 | 0.21 | 21 |
| Djibouti | 1 | 12.9 | 1.8 | 3.23 | 2.98 - 3.48 | 0.03 | 108 |
| Fiji | 0.9 | 9.8 | 5.7 | 3.21 | 2.97 - 3.45 | 0.06 | 54 |
| Guyana | 0.79 | 1.8 | 13.5 | 3.06 | 2.84 - 3.28 | 0.13 | 24 |
| Bhutan | 0.78 | 2.5 | 13.3 | 3.11 | 2.89 - 3.33 | 0 | NA |
| Luxembourg | 0.64 | 12.6 | 3.1 | 2.15 | 1.99 - 2.32 | 0.08 | 27 |
| Suriname | 0.59 | 2.4 | 12.6 | 2.23 | 2.07 - 2.38 | 0.15 | 15 |
| Cape Verde | 0.56 | 3.7 | 10 | 1.91 | 1.77 - 2.04 | 0.06 | 32 |
| Malta | 0.44 | 11.1 | 4.6 | 1.51 | 1.39 - 1.63 | 0.06 | 25 |
| Brunei | 0.44 | 15.7 | 0 | 1.46 | 1.35 - 1.58 | 0 | NA |
| Belize | 0.4 | 1.4 | 14 | 1.6 | 1.49 - 1.72 | 0.07 | 23 |
| Bahamas | 0.4 | 2.3 | 13 | 1.52 | 1.41 - 1.63 | 0.08 | 19 |
| Iceland | 0.34 | 14.6 | 0.6 | 1.09 | 01.01.2017 | 0 | NA |
| Vanuatu | 0.31 | 11.4 | 3.5 | 1.06 | 0.98 - 1.14 | 0 | NA |
| Barbados | 0.29 | 7.2 | 8 | 1 | 0.93 - 1.08 | 0.01 | 100 |
| Aruba | 0.11 | 11.2 | 4.2 | 0.36 | 0.33 - 0.39 | 0.03 | 12 |
| Seychelles | 0.1 | 10 | 5.1 | 0.34 | 0.31 - 0.36 | 0.02 | 17 |
